# Heterologous prime-boost immunization with CoronaVac and Convidecia

**DOI:** 10.1101/2021.09.03.21263062

**Authors:** Jingxin Li, Lihua Hou, Xiling Guo, Pengfei Jin, Shipo Wu, Jiahong Zhu, Hongxing Pan, Xue Wang, Zhizhou Song, Jingxuan Wan, Lunbiao Cui, Junqiang Li, Xuewen Wang, Lairun Jin, Jingxian Liu, Fengjuan Shi, Xiaoyu Xu, Yin Chen, Tao Zhu, Wei Chen, Fengcai Zhu

## Abstract

**Background:** The safety and immunogenicity of heterologous prime-boost COVID-19 vaccine regimens with one shot of a recombinant adenovirus type-5-vectored COVID-19 vaccine Convidecia has not been reported.

**Methods:** We conducted a randomized, controlled, observer-blinded trial of heterologous prime-boost immunization with CoronaVac and Convidecia in healthy adults 18-59 years of age. Eligible participants who were primed with one or two doses of CoronaVac were randomly assigned at a 1:1 ratio to receive a booster dose of Convidecia or CoronaVac. Participants were masked to the vaccine received but not to the three-dose or two-dose regimen. The occurrences of adverse reactions within 28 days after the vaccination were documented. The geometric mean titers of neutralizing antibodies against live SARS-CoV-2 virus were measured at 14 and 28 days after the booster vaccination.

**Results:** Between May 25 and 26, 2021, a total of 300 participants were enrolled. Participants who received a booster shot with a heterologous dose of Convidecia reported increased frequencies of solicited injection-site reactions than did those received a homogeneous dose of CoronaVac, but frequencies of systemic reactions. The adverse reactions were generally mild to moderate. The heterologous immunization with Convidecia induced higher live viral neutralizing antibodies than did the homogeneous immunization with CoronaVac (197.4[167.7, 232.4] vs. 33.6[28.3, 39.8] and 54.4[37. 9, 78.0] vs. 12.8[9.3, 17.5]) at day 14 in the three- and two-dose regimen cohort, respectively.

**Conclusions:** The heterologous prime-boost regimen with Convidecia after the priming with CoronaVac was safe and significantly immunogenic than a homogeneous boost with CoronaVac (ClinicalTrials.gov, number NCT04892459).

## Introduction

The coronavirus disease 2019 (COVID-19) pandemic, which is caused by the infection of severe acute respiratory syndrome coronavirus 2 (SARS-CoV-2), has imposed an enormous disease burden worldwide.^1-3^ The mass vaccination campaigns of effective COVID-19 vaccines are considered as crucial measures to control the pandemic.^4,5^

Currently, more than 15 vaccines have been granted by national regulatory authorities for emergency use, among which six vaccines have been listed for WHO Emergency Use Listing, including adenovirus-based ChAdOx1 nCoV-19 (AstraZeneca), Ad26.COV2.S (Johnson & Johnson), mRNA-based BNT162b2 (BioNTech/Pfizer), mRNA-1273 COVID-19 vaccine (Moderna), and two inactivated vaccines Sinopharm (Beijing institute) and CoronaVac (Sinovac) developed in China.^6^

Since March, 2021, China has begun a national mass vaccination campaign with the vaccine CoronaVac and Sinopharm, as well as other national authorized inactivated COVID-19 vaccines and a recombinant adenovirus type-5-vectored COVID-19 vaccine Convidecia (CanSino). According to the results from phase 3 trials, CoronaVac showed a 50.4% of vaccine efficacy against COVID-19 diseases in Brazil,^7^ and one shot of Convidecia had about 65% of efficacy in preventing symptomatic diseases.^8^ Both of the vaccines were found to be immunogenic and effective, but there is a concern about the waning of vaccine-elicited neutralizing antibodies might make the vaccine inadequately protective in a longer period. Besides, the SARS-CoV-2 variants, with an increased infectivity and transmissibility, may partially escape from vaccine-elicited neutralizing antibodies, and lead to a further decrease of vaccine protection against COVID-19 diseases.

Although, the justification and necessity of a booster vaccination of COVID-19 vaccine is still disputed, clinical trials with heterologous or homogeneous additional dose after the priming series are performed. Heterologous schedules incorporating COVID-19 vaccines across different platforms showed a superior immunogenicity to heterologous schedules in animal studies.^9^ Theoretically, heterologous schedules may promote the maturation of antibody affinity and influence the breadth of immunization by inserting different antigens, types of vectors, delivery routes, doses, or adjuvants at different times.^10^ There are dozens of various ongoing heterologous booster studies after the primary series, but most of them focused on the ChAdOx1 nCoV-19, BNT162b2 and mRNA-1273.^11^ None was heterologous boost study after priming with inactivated COVID-19 vaccines.

Up to now, in mainland China, more than 2.0 billion doses of COVID-19 vaccines have been administered in population, and over 50% of which were inactivated vaccine CoronaVac.^12^ Besides, CoronaVac has been authorized to use in 39 countries or areas and been deployed globally, including Brazil, Malaysia, Mexico, Pakistan, Chile, Egypt, Indonesia, Nepal, Turkey. While, Convidecia has been authorized in eight countries or areas.^13^

Here, we reported the safety and immunogenicity of the first heterologous prime-boost immunization with an inactivated SARS-CoV-2 vaccine (CoronaVac) and a recombinant adenovirus type-5-vectored COVID-19 vaccine (Convidecia) in Chinese adults at 18 to 59 years of age.

## Methods

### Objectives, Participants, and Oversight

We conducted a randomized, controlled, observer-blinded trial to access the safety and immunogenicity of heterologous prime-boost immunization with CoronaVac and Convidecia. Healthy participants, male or female, aged between 18 and 59 years, who have received one dose of CoronaVac in the past 1∼3 months or two doses of CoronaVac in the past 3∼6 months were recruited for screening of eligibility. Participants with a previous clinical or virologic COVID-19 diagnosis or SARS-CoV-2 infection, and women with positive urine pregnancy test results were excluded from this study. The occurrence of adverse reactions and immune responses in eligible participants who were administrated with heterologous Convidecia were compared with those received a homogeneous third dose of CoronaVac.

This was a proof of concept trial, initiated by investigators from Jiangsu Provincial Center of Disease Control and Prevention. The trial protocol was reviewed and approved by the institutional review board of the Jiangsu Provincial Center of Disease Control and Prevention, and no protocol change was made after the initial of the study. This trial was conducted following the principles of the Declaration of Helsinki and local guidelines.

### Procedures

We recruited participants form one clinic site in Lianshui County, Jiangsu Province. Eligible participants who competed the prime vaccination of two doses of CoronaVac in the past 3∼6 months were randomly assigned at a 1:1 ratio to receive a booster dose of Convidecia (group A, heterologous boost dose) or CoronaVac (group B, homogeneous boost dose). While, participants who were primed with one dose of CoronaVac in the past 1∼2 months were randomized in a 1:1 ratio to receive a second dose of Convidecia (group C, heterologous dose) or CoronaVac (group D, homogeneous dose). The vaccination records were verified by investigators through the electronic registration system for COVID-19 vaccine immunization. An interactive Web-based response system was used for randomization, and the randomization lists were generated by an independent statistician using SAS (version 9.4).

We masked participants, investigators, laboratory staff, and outcome assessors to the allocation of treatment groups, but not to the three-dose or two-dose regimen. Designated unblinded personnel were responsible for the preparation and administration of the vaccination, and were forbidden to reveal the identity of the study vaccines to the participants or other investigators. All the vaccinations were administered intramuscularly.

### Safety

The primary endpoint for safety objective was the occurrence of adverse reactions within 28 days after the vaccination. Participants were observed at the clinic for 30 minutes after the vaccination for any immediate vaccine-associated reactions, and then were instructed to keep a daily record of any solicited or unsolicited adverse events for the next 14 days. Solicited injection-site events included pain, redness, swelling, induration, itch and cellulitis, while systemic events included fever, malaise, muscle ache, joint pain, fatigue, nausea, headache and so on. Unsolicited adverse events within 28 days reported by the participants were also collected. Severity of adverse events are graded according to the standard guidelines issued by the China State Food and Drug Administration, and the causality with immunization before unmasking. Serious adverse events self-reported by participants were documented throughout the study.

### Immunogenicity

Participants donated blood samples at baseline before receiving the booster dose, and at 14 and 28 days after receiving the booster dose. The primary endpoint for immunogenicity was the geometric mean titres (GMTs) of neutralizing antibodies against live SARS-CoV-2 virus at 14 days after the booster vaccination. Live viral neutralizing antibody titer in serum was determined by using a cytopathic effect (CPE)-based microneutralization assay with a wild-type SARS-CoV-2 virus strain BetaCoV/Jiangsu/JS02/2020 (EPI_ISL_411952) in Vero-E6 cells. Serum dilutions were then mixed with the same volume of virus solution to achieve a 100 TCID_50_ (50% tissue culture infectious dose) per well. The reported titer was the reciprocal of the highest sample dilution that protected at least 50% of cells from CPE. The serum dilution for microneutralization assay started from 1:4, and the seropositivity was defined as the titer ≥1:4.

Receptor binding domain (RBD)- and N-specific ELISA antibody responses were measured at the same time points, using an indirect ELISA assay. RBD-binding IgG isotype in serum was also determined by ELISA, and the type 1 helper T cells (Th1)-dependent IgG1 vs. type 2 helper T cells (Th2)-dependent IgG4 antibody subclasses were calculated to evaluate the Th1/Th2 profiling. The WHO international standard for anti-SARS-CoV-2 immunoglobulin (NIBSC code 20/136) was used side by side as reference with the serum samples measured in this study for calibration and harmonization of the serological assays.^14^ The conversion factors to international standard units were showed in the appendix 1. Fold increase of antibody responses and types of binding antibodies IgG against SARS-CoV-2 S protein in serum post-vaccination in each treatment group were also analyzed. To evaluate cellular immunity, we isolated peripheral blood mononuclear cells (PBMCs) from blood samples of the first 50 and 30 participants in the three-dose regimen and two-dose regimen cohort at days 14 post the boost, respectively. PBMCs were stimulated with a SARS-CoV-2 spike peptide-pool and cytokine secretions of Th1 (interferon-gamma [IFN-γ], and tumor necrosis factor-α [TNFα]), and Th2 (IL-4, IL-5 and IL-13) were determined by ELISpot.

### Statistical Analysis

The sample size calculation was based on the hypothesis on a boost vaccination following the two-dose of inactivated vaccine regimen, and performed by using Power Analysis and Sample Size (PASS) software. We assumed that the GMT of neutralizing antibodies was about 1:40 at baseline before receiving the booster immunization (i.e. three to six months after receiving two doses of inactivated vaccines). After the boost vaccination, the GMTs were expected to reach 1:80 for those receiving a homogeneous dose of CoronaVac, and 1:160 for those receiving a or a heterogeneous of Convidecia at days 14. Equal standard deviation of GMTs of 4 was estimated for both groups. A sample size of 100 per treatment group would provide at least 90% power to detect a difference in log-transformed postvaccination GMTs at the one-sided 2.5% significance level. In this study, a heterogeneous vaccination following one dose of inactivated vaccine were also explored with 50 persons per group, for which the power was not calculated.

We assessed the number and proportion of participants with adverse reactions post vaccination. The antibodies against SARS-CoV-2 were presented as GMTs, geometric mean fold increases (GMFIs) and the proportion of participants with at least four-fold increase with 95% CIs. The cellular responses were shown as the average number of positive cells per million peripheral blood mononuclear cells (PBMCs). We used the χ^2^ test or Fisher’s exact test to analyze categorical data, T test to analyze the log transformed antibody titers, and Wilcoxon rank-sum test for non-normal distributed data. The correlation between concentrations of log-transformed neutralizing antibodies and binding antibody was analyzed using Pearson’s correlation.

The primary analysis was performed based on the intervention modified intention-to-treat cohort. Statistical analyses were done by using SAS (version 9.4) or GraphPad Prism 8.0.1. This study is registered with ClinicalTrials.gov, number NCT04892459.

## Results

### Participants

Between May 25 and 26, 2021, we recruited 302 participants for screening. A total of 300 participants were enrolled, of whom 200 primed with two doses of inactivated vaccine CoronaVac, and 100 primed with one dose of CoronaVac were randomized (figure 1). One participant only received one prime dose, but was wrongly classified into the cohort having two doses primed, and then randomized to receive a heterologous boost dose of Convidecia. We reclassified this participant into group C. Two participants who were randomized to group A but were wrongly administrated with a homogeneous boost dose of CoronaVac, were classified into group B. 299 of these participants received a booster dose on day 0, and completed seven days follow-up to assess safety. We obtained serum samples from 298 participants at day 14, and 293 participants at day 28. The demographic characteristics of participants are shown in table 1. Approximately 11.8∼27.1% of the participants who completed two doses in the last 3 to 6 months and 4.0∼5.9% of those who received one dose in the last 1 to 2 months, showed positive neutralizing antibody against SARS-CoV-2 in serum at the enrollment before receiving a boost vaccination.

**Table 1.**
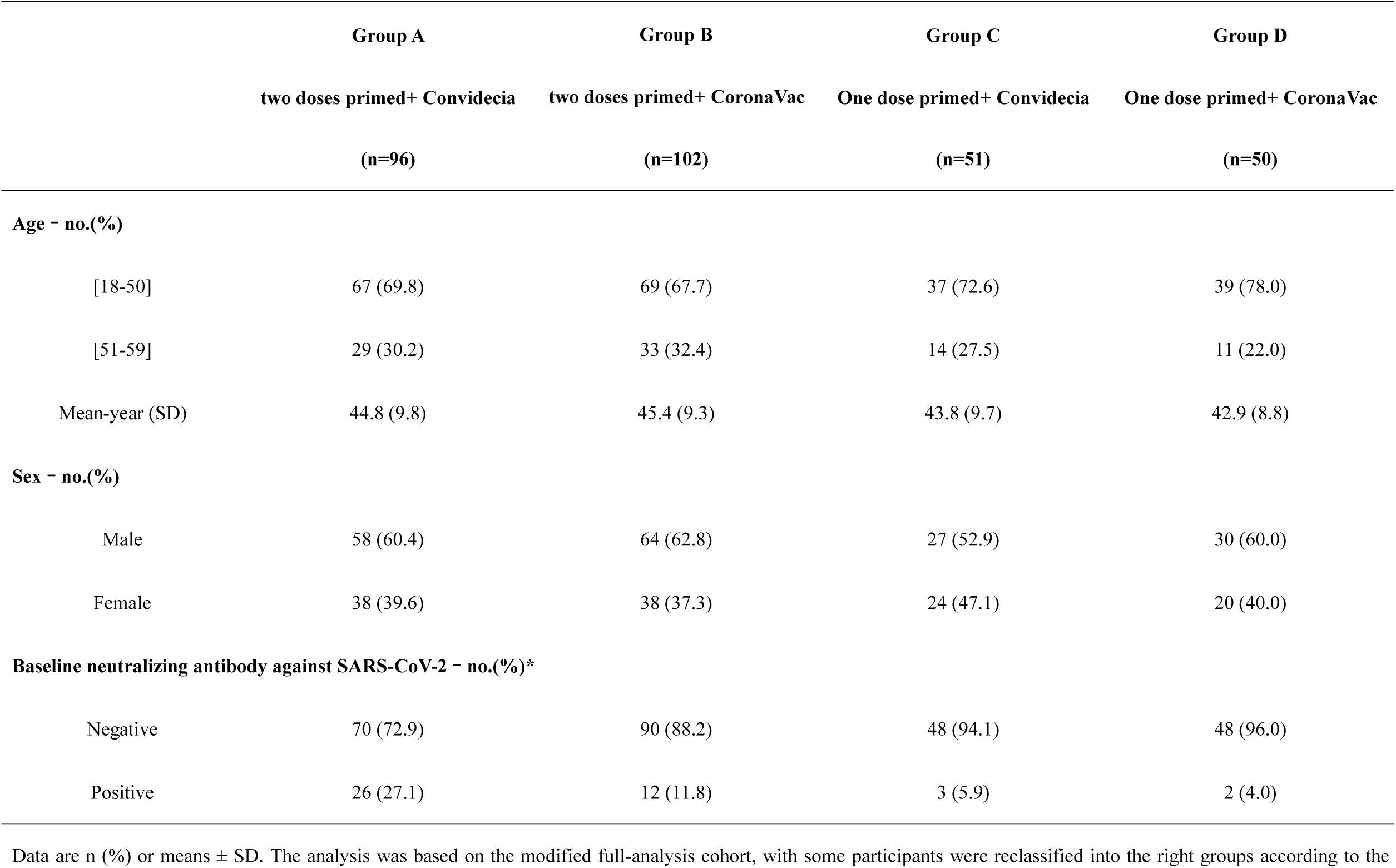
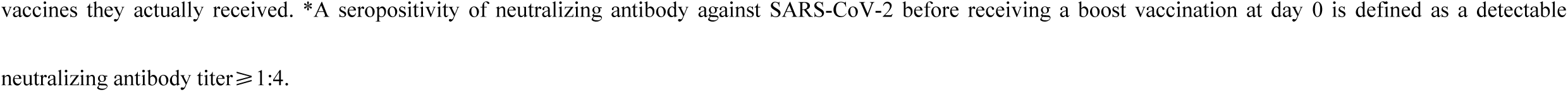
Baseline characteristics of the participants in the modified full-analysis cohort.

|  | **Group A** | **Group B** | **Group C** | **Group D** |
| --- | --- | --- | --- | --- |
|  | **two doses primed+ Convidecia** | **two doses primed+ CoronaVac** | **One dose primed+ Convidecia** | **One dose primed+ CoronaVac** |
|  | **(n=96)** | **(n=102)** | **(n=51)** | **(n=50)** |
| **Age - no.(%)** |  |  |  |  |
| [18-50] | 67 (69.8) | 69 (67.7) | 37 (72.6) | 39 (78.0) |
| [51-59] | 29 (30.2) | 33 (32.4) | 14 (27.5) | 11 (22.0) |
| Mean-year (SD) | 44.8 (9.8) | 45.4 (9.3) | 43.8 (9.7) | 42.9 (8.8) |
| **Sex - no.(%)** |  |  |  |  |
| Male | 58 (60.4) | 64 (62.8) | 27 (52.9) | 30 (60.0) |
| Female | 38 (39.6) | 38 (37.3) | 24 (47.1) | 20 (40.0) |
| **Baseline neutralizing antibody against SARS-CoV-2 - no.(%)\*** |  |  |  |  |
| Negative | 70 (72.9) | 90 (88.2) | 48 (94.1) | 48 (96.0) |
| Positive | 26 (27.1) | 12 (11.8) | 3 (5.9) | 2 (4.0) |

Data are n (%) or means ± SD. The analysis was based on the modified full-analysis cohort, with some participants were reclassified into the right groups according to the
vaccines they actually received. \*A seropositivity of neutralizing antibody against SARS-CoV-2 before receiving a boost vaccination at day 0 is defined as a detectable neutralizing antibody titer $\geq 1:4$ .

**Figure 1.**
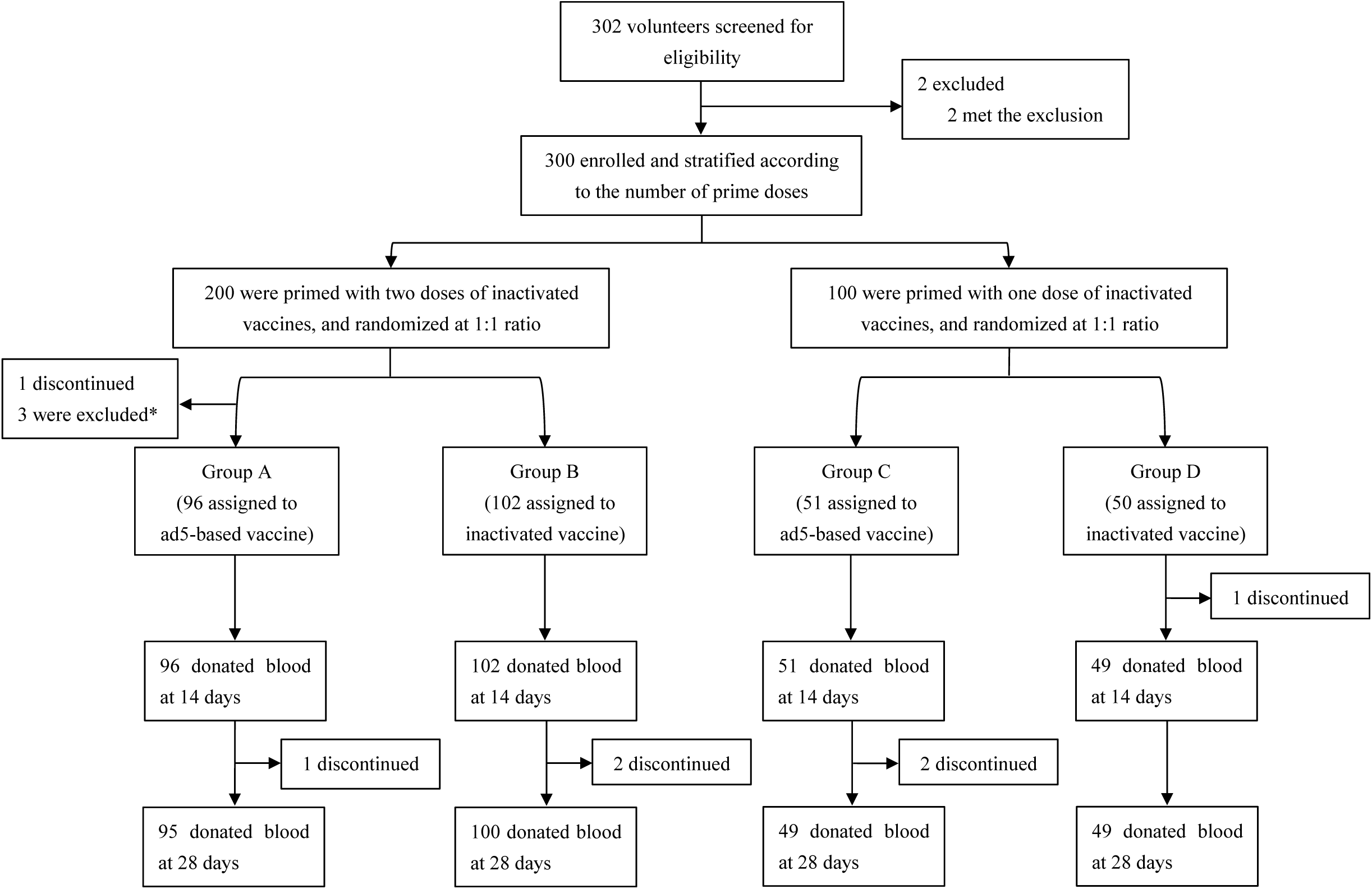
Trial profile. *2 participants were randomized to group A, but were wrongly administrated with an inactivated vaccine and then classified to group B. 1 participant was only primed one dose, but was wrongly classified into the population with two doses prime vaccination, and randomized to group A to receive one dose of ad5-based vaccine. We reclassified this participant to Group C.

### Safety

In both two-dose and three-dose regimen cohorts, Convidecia recipients reported more adverse reactions than CoronaVac recipients with p values of <0.001 and 0.019, respectively (Table 2). Participants in the two-dose regimen cohort, who received the second dose with a heterologous dose of Convidecia reported significant higher occurrence of solicited injection-site reactions than did those received a homogeneous dose of CoronaVac. While, the systemic reactions were reported at similar frequencies between the two groups. In the three-dose regimen cohort, participants received the third dose with a Convidecia following a heterologous prime-boost immunization had significant more solicited injection-site and systemic reactions than those received a homogeneous dose of CoronaVac (29.2% vs. 2.9%, p<0.001, and 13.5% vs. 2.9%, p=0.006). The injection-site and systemic reactions were generally mild to moderate in severity, and typically resolved within 1 or 2 days. In both regimen cohorts, injection-site pain was the most common injection-site reaction. Severe injection-site pain after Convidecia was reported in 2.1% of the participants, while no severe pain was reported after the administration of CoronaVac. Fever and fatigue were the most frequently reported systemic reactions. Unsolicited adverse reactions within 28 days after the vaccination had a low incidence in both treatment groups in two-dose or three-dose regimen cohort. No severe fever was noted, but the use of antipyretic agents was slightly more frequent among Convidecia recipients than among those who received CoronaVac (4.8% vs. 0.7%). No thromboses or vaccine-related anaphylaxis, or serious adverse event was seen in any cohort of the participants.

**Table 2.**
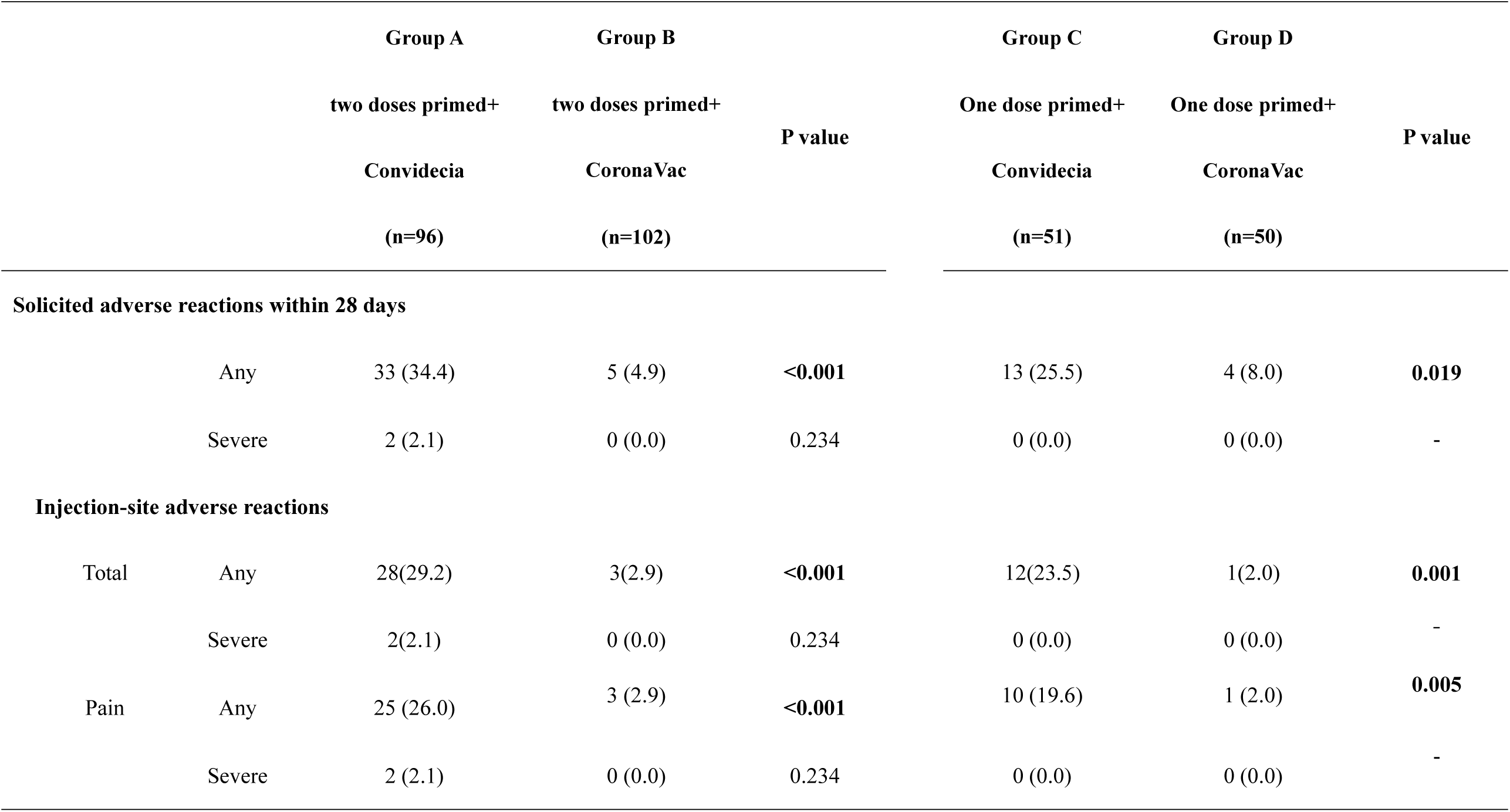
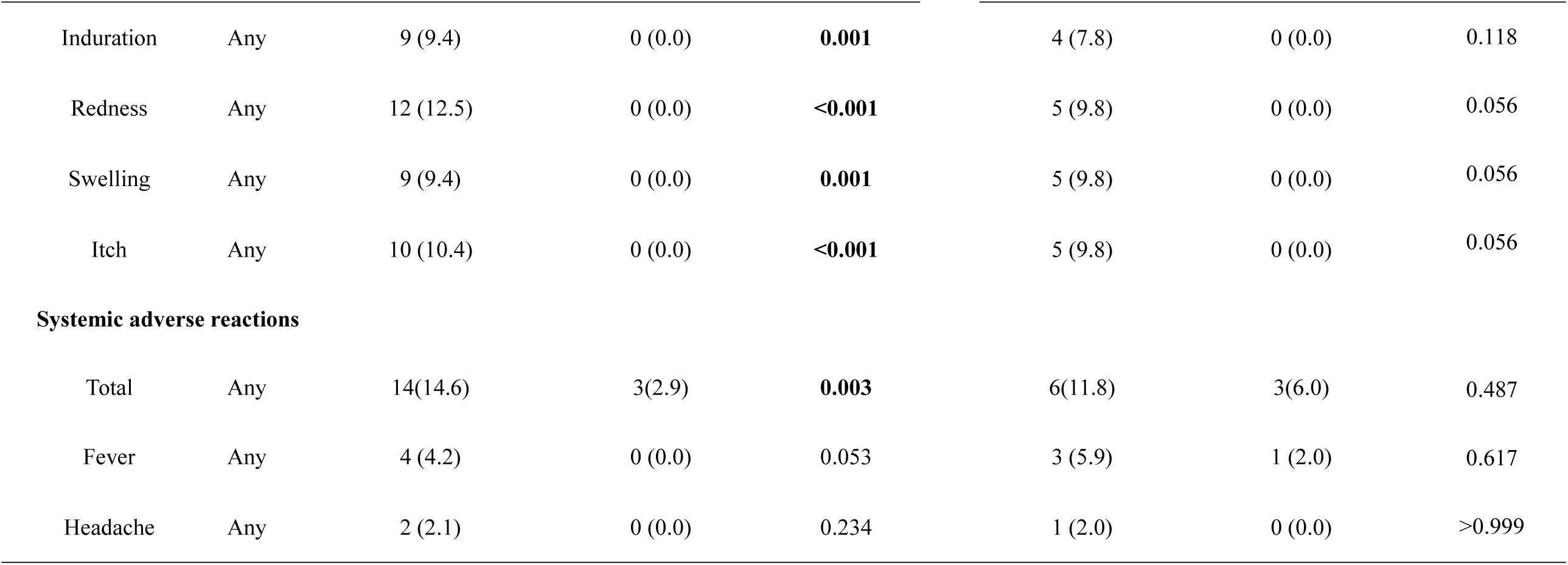
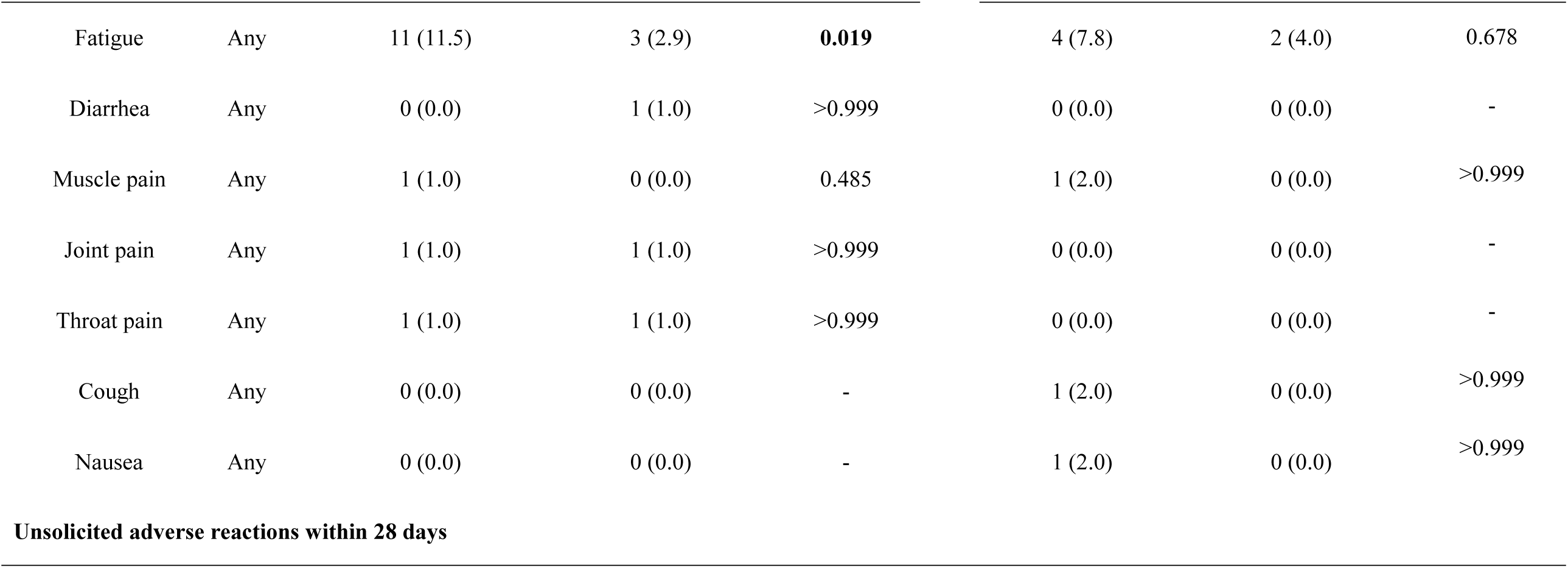
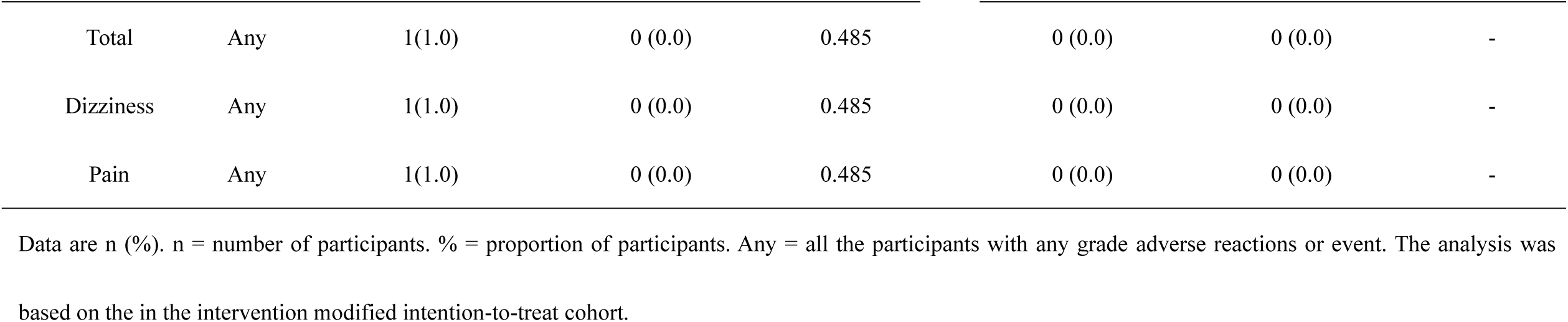
Solicited and unsolicited adverse reactions occurred within 28 days after the vaccination.

### Immunogenicity

Significant responses were observed after the booster dose vaccination in both three-dose and two-dose cohort (table 3). GMTs (PRNT_50_) of neutralizing antibodies at day 0 before the vaccination were about 2.5 (95%CI 2.3, 2.7) and 2.2 (2.1, 2.3) in the three-dose regimen cohort, increasing to 197.4 (167.7, 232.4) and 33.6 (28.3, 39.8) at day 14 after receiving Convidecia and CoronaVac, respectively (<0.0001). While, PRNT_50_ neutralizing antibodies GMTs of 2.1 (95%CI 2.0, 2.3) and 2.1 (2.0, 2.1) at day 0 before the vaccination were noted in the two-dose regimen cohort, which increased to 54.4 (37. 9, 78) and 12.8 (9.3, 17.5) at day 14 after receiving Convidecia and CoronaVac as the second dose, respectively (<0.0001). The heterologous boost with Convidecia demonstrated an at least 78-fold increase in the average neutralizing antibody levels, and the homogeneous boost with CoronaVac did 15.2-fold increase, for the three-dose regimen group post the boost. In the two-dose regimen, neutralizing antibody GMTs in participants receiving heterologous boost with Convidecia increased at least 25.7-fold, while those receiving homogeneous boost with CoronaVac did 6.2-fold increase. At day 28 post the boost, the average neutralizing antibody levels decreased, but still heterologous boost group showed significant higher neutralizing antibody GMTs than did the homogeneous boost group (150.3 [128.6, 175.7] vs. 35.3 [29.4, 42.4] in the three-dose regimen cohort, and 49.6 [35.1, 70.2] vs. 10.6 [8.3, 13.5] in the two-dose regimen cohort).

**Table 3.** GMT, seroconversion rate, and GMFI of neutralizing antibodies to live SARS-CoV-2 before and after a heterogeneous or homologous boost vaccination.

|  | Group A | Group B |  | Group C | Group D |  |
| --- | --- | --- | --- | --- | --- | --- |
|  | two doses primed+ | two doses primed+ |  | one dose primed+ | one dose primed+ |  |
|  | Convidecia | CoronaVac | *P* value | Convidecia | CoronaVac | *P* value |
|  | (n=96) | (n=102) |  | (n=51) | (n=50) |  |
| Day 0 |  |  |  |  |  |  |
| GMT | 2.5 (2.3, 2.7) | 2.2 (2.1, 2.3) | 0.0119 | 2.1 (2.0, 2.3) | 2.1 (2.0, 2.1) | 0.4876 |
| Day 14 |  |  |  |  |  |  |
| GMT | 197.4 (167.7, 232.4) | 33.6 (28.3, 39.8) | <0.0001 | 54.4 (37. 9, 78) | 12.8 (9.3, 17.5) | <0.0001 |
| Seroconversion | 100.0 (1.0, 1.0) | 100.0 (1.0, 1.0) | - | 98.0 (0.9, 1.0) | 87.8 (0.8, 1.0) | 0.0572 |
| GMFI | 78.3 (66.4, 92.4) | 15.2 (12.8, 17.9) | <0.0001 | 25.7 (18.0, 36.9) | 6.2 (4.6, 8.4) | <0.0001 |
| Day 28 |  |  |  |  |  |  |
| GMT | 150.3 (128.6, 175.7) | 35.3 (29.4, 42.4) | <0.0001 | 49.6 (35.1, 70.2) | 10.6 (8.3, 13.5) | <0.0001 |
| Seroconversion | 100.0 (1.0, 1.0) | 99.0 (0.9, 1.0) | 1.0000 | 100.0 (0.9, 1.0) | 93.9 (0.8, 1.0) | 0.2423 |
| GMFI | 59.5 (50.9, 69.6) | 15.9 (13.2, 19.1) | <0.0001 | 23.4 (16.6, 33.1) | 5.2 (4.1, 6.5) | <0.0001 |

Data are GMT (95% CI), number of participants (%; 95%CI), or GMFI (95% CI). n= the number of participants included the intervention modified intention-to-treat cohort. The *P* values are the results of comparison between the two treatment groups. Measurements were on day 0 were taken immediately before vaccination. GMT=geometric mean titre. GMFI=geometric mean fold increase.

In line with the neutralizing antibody titers, both heterologous and homogeneous boost could induce significant immunological memory responses with a large enhancement of RBD-binding IgG (figure 2A). But the heterologous boost with Convidecia elicited higher levels of RBD-binding IgG GMTs than did homogeneous boost with CoronaVac (3090.1 [95%CI 2636.1, 3622.3] vs. 369.0 [95%CI 304.2, 447.5] in the three-dose regimen cohort, and 941.8 (663.9, 1336.1) vs. 154.1 (116.3, 204.3) in the two-dose regimen cohort) at day 14 (appendix 2). The anti-RBD IgG isotype showed that the predominant binding antibody responses were associated with IgG1 after the boost in both heterologous or homogeneous vaccine groups (appendix 3). Very mild responses from IgG3 were found in participants receiving heterologous Convidecia, but not in those receiving homogeneous CoronaVac. Nearly no responses of IgG2 and IgG4 were observed across the treatment groups. At day 14, the mean ratios of IgG1/IgG4 were 42.4 (95%CI 35.6, 50.6) vs. 6.1 (5.2, 7.1), and 24.4 (17.7, 33.6) vs. 3.8 (3.1, 4.6) for the Convidecia vs. CoronaVac groups in the three-dose and two-dose regimen cohorts, respectively. Participants in both the cohorts had similar N-specified binding antibodies at day 0 before the vaccination, but only participants who received CoronaVac showed increases of IgG titers to both RBD and N protein after the boost dose (figure 2B). Convidecia recipients showed no increase of N-specified binding antibodies post the boost. The association between the RBD-binding antibodies and neutralising antibody titres against live virus showed a positive correlation coefficient of 0.56∼0.77 post vaccination in the heterologous and homogeneous prime-boost groups, respectively (appendix 4).

**Figure 2.**
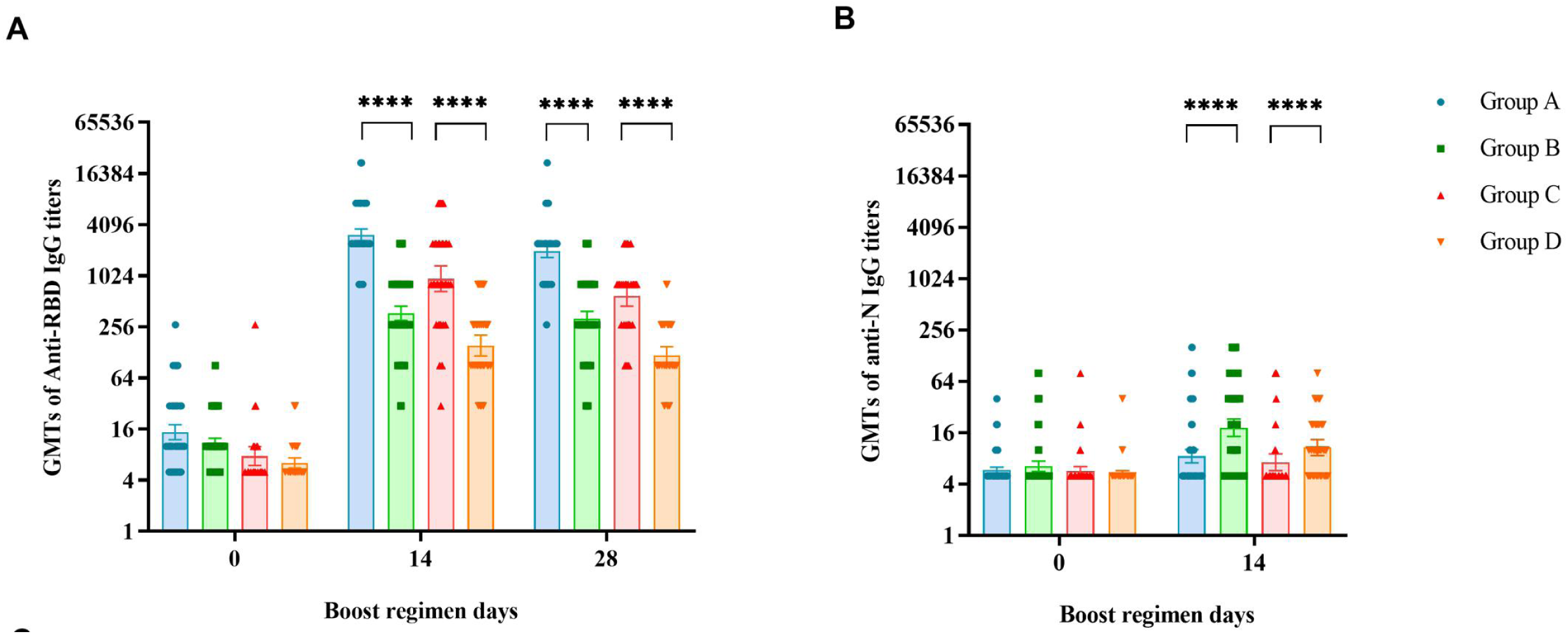
Receptor binding domain (RBD)-, N-specific ELISA antibody responses before and after a heterogeneous or homologous boost vaccination. Group A: primed with two doses of CoronaVac + Convidecia; Group B: primed with two doses of CoronaVac + CoronaVac; Group C: primed with one dose of CoronaVac + Convidecia; Group D: primed with one dose of CoronaVac + CoronaVac. ****p value <0.0001.

We observed a profound increase in the levels of Th1-biased cytokine IFN-γ compared with the pre-boost across the treatment groups in both cohorts, at 14 days after the boost dose (figure 3A). Participants who were in the three-dose regimen cohort, had medians of IFN-γ+ spot counts of 100 per 10^6^ PBMCs (IQR 60, 165) and 90 per 10^6^ PBMCs (40, 230) after receiving heterologous Convidecia, and homogeneous CoronaVac, respectively. In the participants in the two-dose regimen cohort, somewhat lower geometric mean IFN-γ+ spot counts of 65 per 10^6^ PBMCs (IQR 45, 95) in the heterologous Convidecia group and 40 per 10^6^ PBMCs (30, 60) in the homogeneous CoronaVac group were noted. However, the pre-boost cytokines TNF-α were relatively high across the groups and no increases of TNF-α were found after the boost (figure 3B). Th2 responses were detected at minimal levels in the heterologous Convidecia groups, as observed by IL-4, IL-5, and IL-13 responses (figure 3C, D, E). Overall, we observed a robust Th1 biased cellular immune response, but low levels of Th2-biased cytokines IL-4, IL-5, and IL-13 in the heterologous vaccination groups, resulting in higher Th1/Th2 cytokine ratios than those in the homogeneous groups (figure 3F).

**Figure 3.**
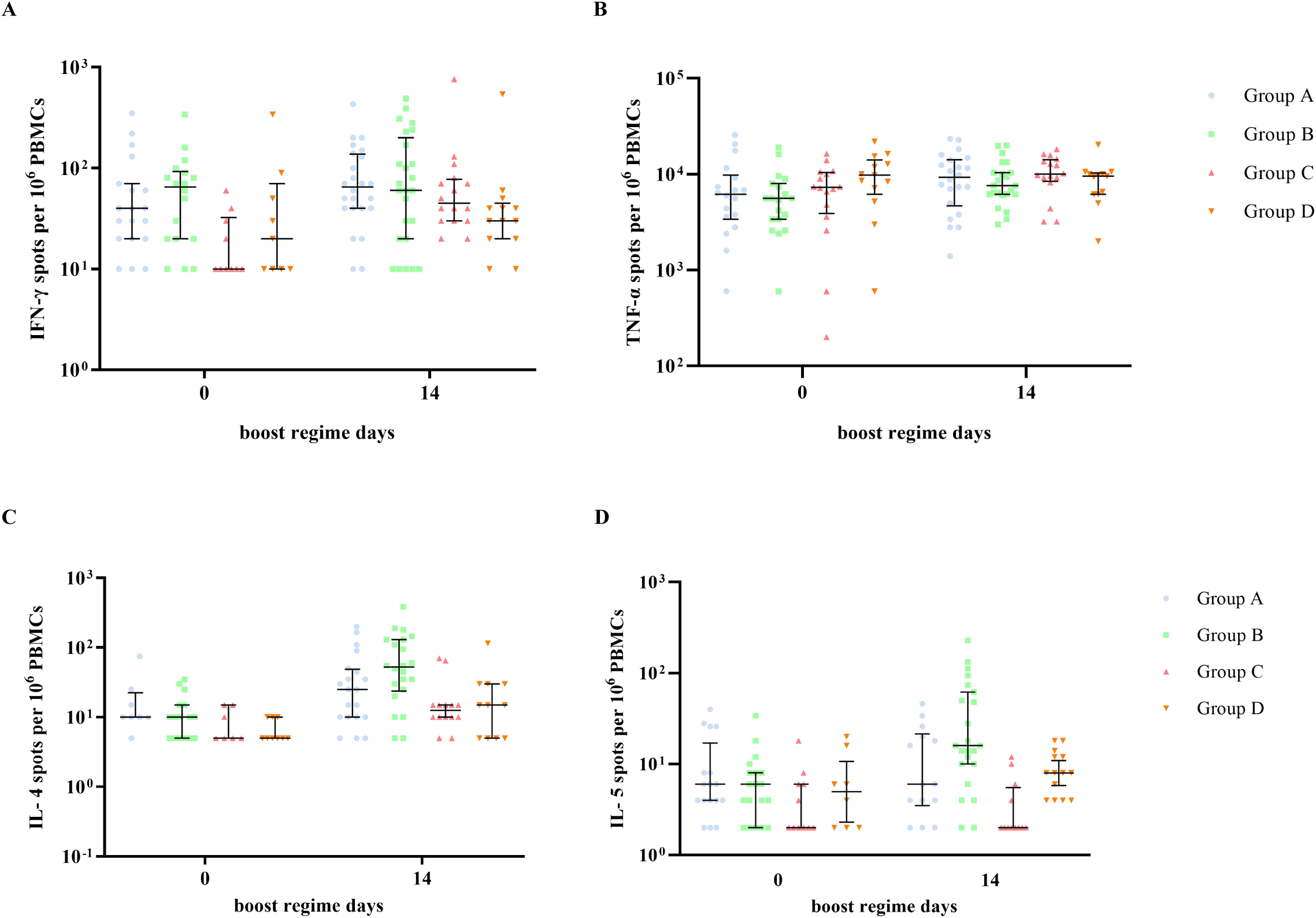
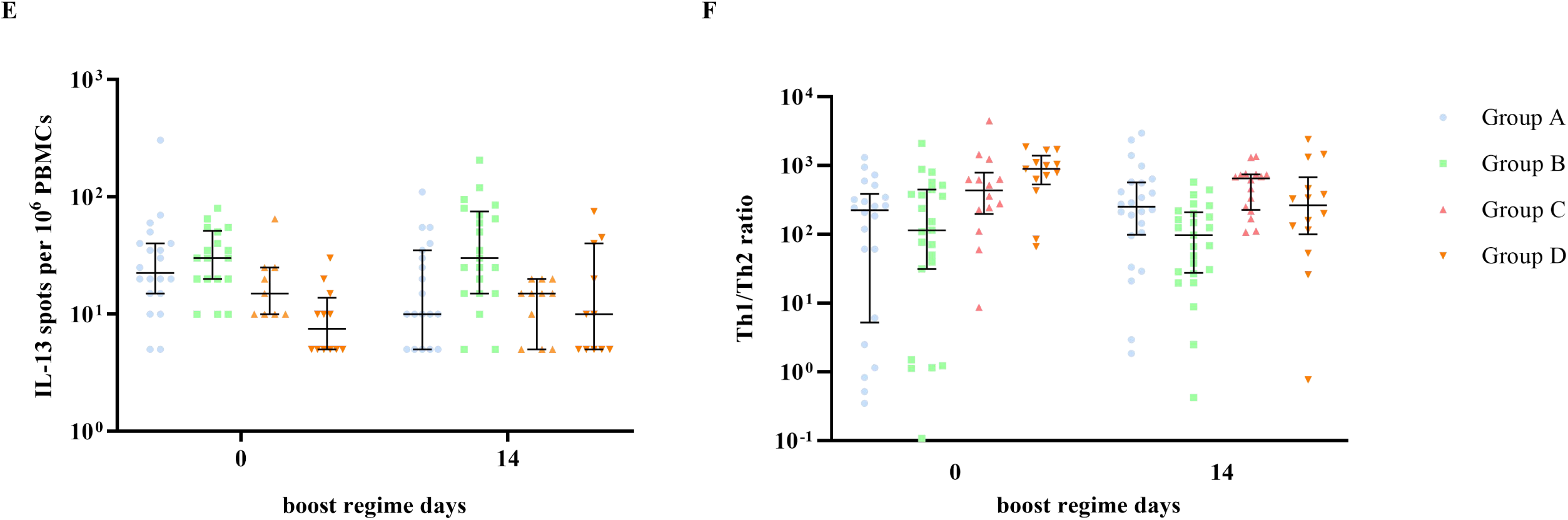
SARS-CoV-2 spike-specific cytokine T cells responses before and after receiving a heterogeneous or homologous boost vaccine. Group A: primed with two doses of CoronaVac + one dose of Convidecia; Group B: primed with two doses of CoronaVac + one dose of CoronaVac; Group C: primed with one dose of CoronaVac + one dose of Convidecia; Group D: primed with one dose of CoronaVac + one dose of CoronaVac. Cytokine T cells were background corrected for unstimulated cells and values lower than 0 were considered negative. Th1/Th2 ratio was calculated by the sum of IFN-γ plus TNF-α cytokine levels divided by the sum of IL-4, and IL-5 plus IL-13 cytokine level. Horizontal bars show the median and error bars show the interquartile range.

## Discussion

Our results suggest that the heterologous prime-boost regimens with one dose of Convidecia administrated at an interval of 1-2 months, or 3-6 months after one- or two-dose of CoronaVac as priming, were safe and highly immunogenic for healthy adults aged 18-59 years. More robust humoral responses and Th1 skewed cellular immune responses were noted following the heterologous boost vaccination of Convidecia, compared with those following the homogeneous boost vaccination of CoronaVac. The peak of neutralizing antibody at day 14 after heterologous boost reached to 197.4 and 54.4 for the three-dose and two-dose regimens, which are equivalent to 616.9 IU/mL and 170.0 IU/mL using the WHO international standard (appendix 1). Assuming that the neutralizing antibody levels correlated with the level of protection for human beings, a heterologous prime-boost vaccination with CoronaVac and Convidecia could be potentially associated with a superior protection to SARS-CoV-2 in the vaccinated human beings. Although slightly higher incidences of injection-site and systemic reactions were found following the heterologous vaccination of Convidecia than those following the homogeneous vaccination of CoronaVac, no severe safety issues were noted.

The concerns about the duration of protection against SARS-CoV-2 induced by the licensed two-dose regimen of inactivated COVID-19 vaccines were raised in both the clinical trials and post-licensure studies, which showed that the humoral immunity waned significantly over time, and protection might be lost, particularly when the variants are dominant.^15-18^ In our study, participants were enrolled within 1-2 months after receiving one dose of inactivated COVID-19 vaccine, or within 3-6 months after completing two doses of inactivated COVID-19 vaccine. We found that the pre-boost antibody titer level of these participants at enrollment were very low, which was in line with that previously reported.^19^ Recently, a study on a booster dose of CoronaVac vaccine in adults aged 18-59 years at 6-month interval induced approximately 3-fold increase of the neutralizing antibody titers above the peak responses induced by two-dose regimen of CoronaVac.^19^ Although homologous prime-boost immunization has typically been effective for the diseases of the Expanded Program of Immunizations, but in our study, heterologous prime-boost are more immunogenic. This may be related to the different natural immune responses activated by the inactivated vaccine and the viral-vectored delivery system expressing only spike protein, which focus the memory responses on the spike and shifts the responses to the inserts rather than the vectors in the immunodominance hierarchy.^20-22^ Our study provided the first evidences on the safety and immunogenicity of a heterologous regimen with the inactivated vaccine and Ad5 vector-based vaccine against SARS-CoV-2 in human being. Although the 28-day homologous two-dose inactivated vaccine regimen was the least immunogenic of the four regimens in our study, this is a licensed vaccine schedule which has hit above the minimum efficacy of 50%, and reduce over 86% of hospitalization and death in both phase 3 trials and post-license studies.^23^

Up to now, at least four studies adopted heterologous prime-boost regimens have been reported. The study of rAd26 and rAd5 vector-based heterologous prime-boost COVID-19 vaccines performed in Russia, inducing a robust immune response and a 91.6% of the efficacy against symptomatic disease.^24,25^ Two heterologous prime-boost vaccination studies with ChAdOx1 nCoV-19 and BNT162b2 elicited higher IgG concentrations than that of a licensed homogeneous schedule (ChAd/ChAd).^11,26^ One study on a heterologous prime-boost vaccination with ChAdOx1 nCoV-19 and mRNA-1273 also demonstrated an increase of serum neutralization titer to the wild-type, and the B.1.351 variant of SARS-CoV-2, in contrast to a heterologous ChAdOx1 nCoV-19 boost.^27^ Only one of the studies reported that a higher reactogenicity was found associated with the heterologous prime-boost vaccination, and others did not.^28^

The first limitation of this study is that only adults aged between 18-59 years were involved in, but not older adults, particularly those over 75 years of age, who are often immunocompromised or with coexisting conditions, and poorly respond to vaccines. We are carrying out another study to evaluate the heterologous prime-boost vaccination with CoronaVac and Convidecia in the older population (NCT04952727). Second, studies on the mechanism of the enhanced immune responses and the detailed B- and T cell activation associated with the heterologous prime-boost were not performed, thus we can only have a speculation about the reasons why this regimen are more immunogenic. Third, we could not access the efficacy of the heterologous prime-boost vaccination regimen against COVID-19 diseases, and the protection of this regimen remain undetermined. But several previous studies found that binding and neutralizing antibodies correlated with COVID-19 risk and were most likely to be able to predict the vaccine efficacy.^29^ Besides, the neutralizing antibody titers against delta variant B.1.617.2 was not reported. As the delta variant has become predominant variants of concern in many countries, whether the heterologous prime-boost regimen could potentially offer an additional protection to the delta variant over currently licensed two-dose inactivated vaccine schedule needs to be answered. At last, the relatively small number of participants in this study was insufficient to identify potential increase of risks for some rare but severe adverse reactions, particularly the immune-mediated events.

In conclusion, the heterologous prime-boost regimen with inactivated vaccine CoronaVac and ad5-vectored vaccine Convidecia were safe and highly immunogenic, increased both humoral and cellular immunity responses significantly, which could be useful for a third dose strategies to be administered to persons who have previously received two doses of inactivated vaccines.

## Data Availability

The results supporting the findings in this study are available upon request from the corresponding authors.

