## Supplementary Appendix for "Heterologous prime-boost immunization with CoronaVac and Convidecia"

**Table of contents**

| **Contents** | **Page** |
| --- | --- |
| **Appendix 1.**  **The conversion factors for values of live SARS-CoV-2 neutralizing antibody assay, and receptor binding domain (RBD)-ELISA assay to WHO international standard units for calibration.** | **2** |
| **Appendix 2.**  **Receptor binding domain (RBD)- or N-specific ELISA antibody responses before and after a heterogeneous or homologous boost vaccination.** | **3** |
| **Appendix 3.**  **Receptor binding domain (RBD)-binding IgG isotypes before and after receiving a heterogeneous or homologous boost vaccine.** | **5** |
| **Appendix 4.**  **Correlations between live SARS-CoV-2 neutralizing antibodies and receptor binding domain (RBD) antibodies by vaccine regimens at day 14 and 28 post-vaccination.** | **6** |

### Appendix 1. The conversion factors for values of live SARS-CoV-2 neutralizing antibody assay, and receptor binding domain (RBD)-ELISA assay to WHO international standard units for calibration.

A. Live SARS-CoV-2 neutralizing antibody assay

A conversion factor of 1/0.32 could be applied to convert the reported results from live SARS-CoV-2 neutralizing antibody titer to “IU/mL”. The WHO reference serum 1000 IU/mL equivalent to live SARS-CoV-2 neutralizing antibody titer of 1:320. The following formula could be used for converting:

Result (IU/mL) = Result (neutralizing antibody titer) / 0.32;

B. Receptor binding domain (RBD)-ELISA assay

A conversion factor of 1/2.43 could be used to convert the specific RBD IgG ELISA antibody titer to “BAU/mL”. The WHO reference serum 1000 BAU/mL equivalent to RBD IgG ELISA antibody titer of 1:2430. The following formula could be used for converting:

Result (BAU/mL) = Result (RBD IgG ELISA antibody titer) / 2.43;

### Appendix 2. Receptor binding domain (RBD)- or N-specific ELISA antibody responses before and after a heterogeneous or homologous boost vaccination.

|  | **Group A**  **two doses primed+ Convidecia  (n=96)** | **Group B**  **two doses primed+ CoronaVac**  **(n=102)** | ***P* value** |  | **Group C**  **one dose primed+ Convidecia  (n=51)** | **Group D**  **one dose primed+ CoronaVac  (n=50)** | ***P* value** |
| --- | --- | --- | --- | --- | --- | --- | --- |
| **Anti-RBD IgG** | | | | | | | |
| Day 0 | | | | | | | |
| GMT | 14.7 (12.0, 18.0) | 10.9 (9.5, 12.5) | **0.0155** |  | 7.7 (6.0, 9.9) | 6.4 (5.6, 7.3) | 0.1940 |
| Day 14 | | | | | | | |
| GMT | 3090.1 (2636.1, 3622.3) | 369.0 (304.2, 447.5) | **<0.0001** |  | 941.8 (663.9, 1336.1) | 154.1 (116.3, 204.3) | **<0.0001** |
| Seroconversion | 100.0 (96.2, 100.0) | 98.0 (93.1,99.5) | 0.4979 |  | 98.0 (89.7, 99.7) | 100.0 (92.7, 100.0) | >0.9999 |
| GMFI | 209.8 (166.4, 264.3) | 33.9 (28.2, 40.7) | **<0.0001** |  | 122.2 (79.1, 188.7) | 24.0 (19.0, 30.2) | **<0.0001** |
| Day 28 | | | | | | | |
| GMT | 1992.1 (1678.4, 2364.5) | 318.4 (259.6, 390.4) | **<0.0001** |  | 591.8 (446.9, 783.6) | 117.8 (92.9, 149.3) | **<0.0001** |
| Seroconversion | 97.9 (92.6, 99.4) | 98.0 (93.0, 99.5) | >0.9999 |  | 95.9 (86.3, 98.9) | 100.0 (92.7, 100.0) | 0.4948 |
| GMFI | 137.3 (105.6, 178.4) | 29.5 (24.3, 35.8) | **<0.0001** |  | 76.5 (53.3,109.8) | 18.3 (14.9, 22.6) | **<0.0001** |
| **Anti-N IgG** | | | | | | | |
| Day 0 | | | | | | | |
| GMT | 6.2 (5.5, 6.9) | 6.2 (5.2, 7.0) | 0.9877 |  | 5.5 (4.9, 6.2) | 5.3 (4.8, 5.8) | 0.5869 |
| Day 14 | | | | | | | |
| GMT | 6.1 (5.5, 6.8) | 26.4 (21.3, 32.8) | **<0.0001** |  | 5.6 (4.9, 6.3) | 12.9 (10.3, 16.1) | **<0.0001** |
| Seroconversion | 1.0 (0.2, 5.7) | 62.8 (53.1, 71.5) | **<0.0001** |  | 0.0 (0.0, 7.0) | 40.8 (28.2, 54.8) | **<0.0001** |
| GMFI | 1.0 (0.9, 1.1) | 4.3 (3.4, 5.4) | **<0.0001** |  | 1.0 (1.0, 1.1) | 2.4 (2.0, 3.0) | **<0.0001** |

Data are GMT (95% CI), number of participants (%, 95%CI), or GMFI (95% CI). n= the number of participants included the intervention modified intention-to-treat cohort. GMT=geometric mean titre. GMFI=geometric mean fold increase. The *P* values are the results of comparison between the two treatment groups (Group A vs. Group B, and Group C vs. Group D). Measurements were on day 0 were taken immediately before vaccination.

### Appendix 3. Receptor binding domain (RBD)-binding IgG isotypes before and after receiving a heterogeneous or homologous boost vaccine.

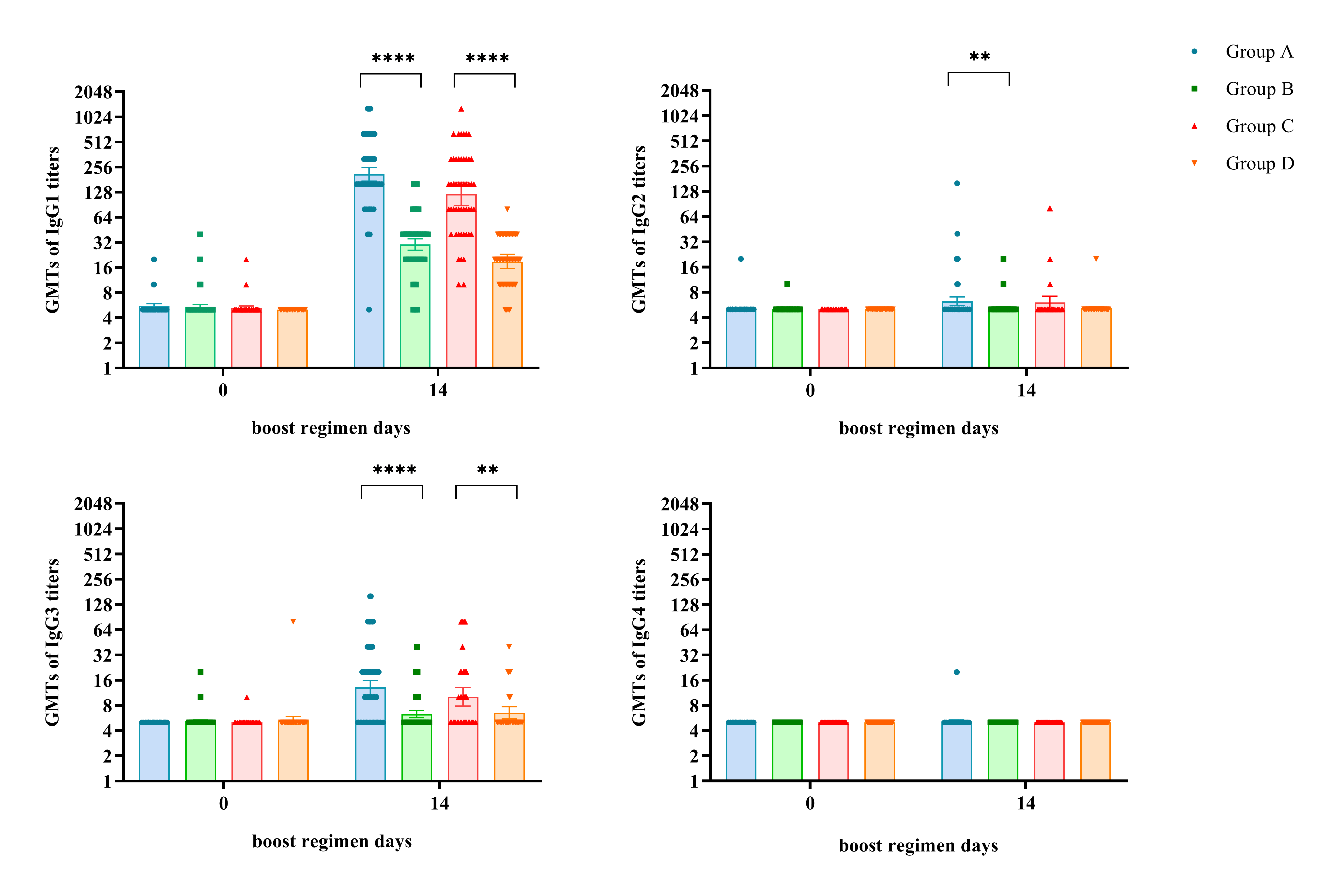

Group A: primed with two doses of CoronaVac + Convidecia; Group B: primed with two doses of CoronaVac + CoronaVac; Group C: primed with one dose of CoronaVac + Convidecia; Group D: primed with one dose of CoronaVac + CoronaVac. **p value<0.01; ****<0.0001.

### Appendix 4. Correlations between live SARS-CoV-2 neutralizing antibodies and receptor binding domain (RBD) antibodies by vaccine regimens at day 14 and 28 post-vaccination.

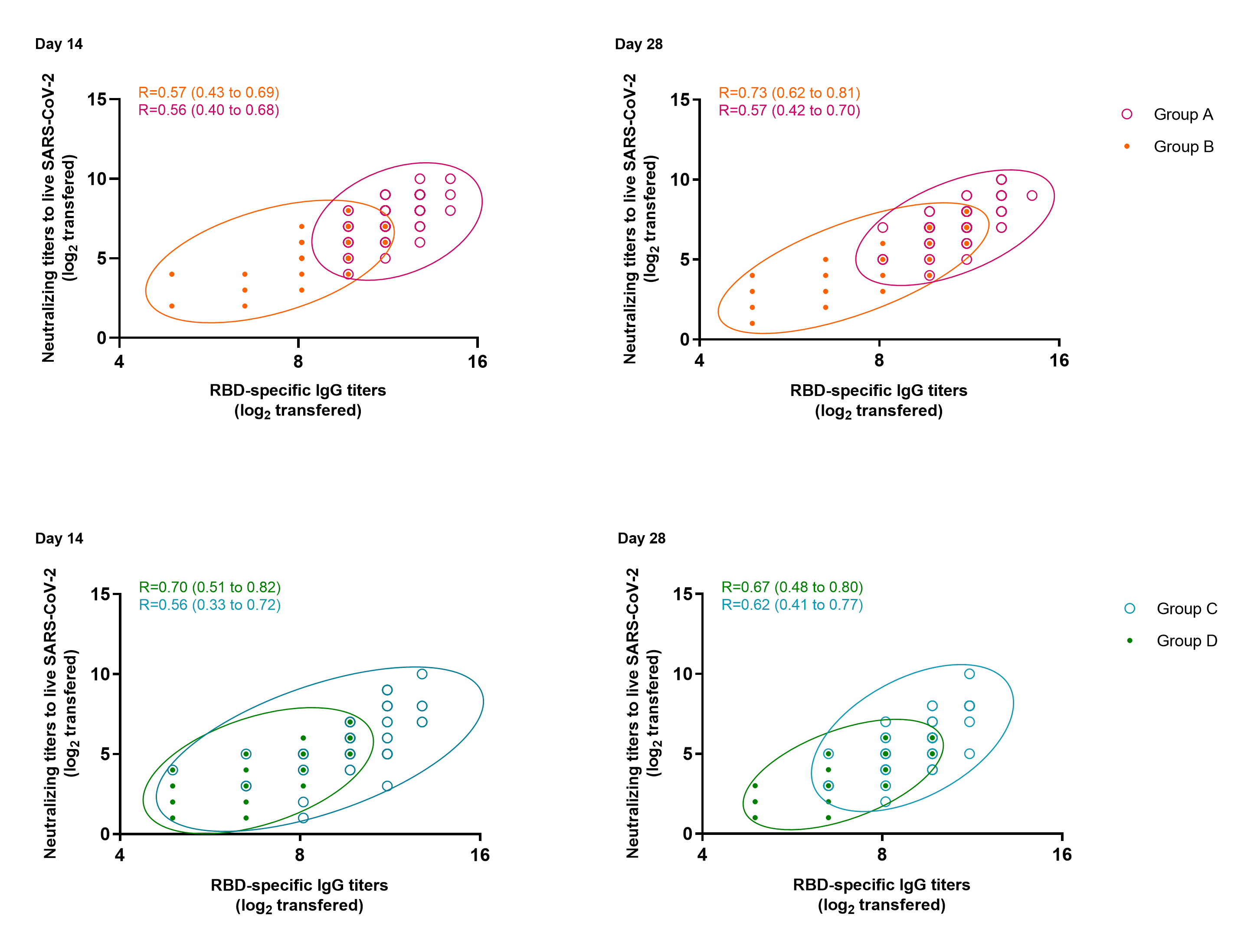

Group A: primed with two doses of CoronaVac + Convidecia; Group B: primed with two doses of CoronaVac + CoronaVac; Group C: primed with one dose of CoronaVac + Convidecia; Group D: primed with one dose of CoronaVac + CoronaVac. Pearson correlation coefficients (95% CIs) are presented for each vaccine schedule.
